# Diversity of CPR manikins for basic life support education: Use of manikin sex, race, and body shape – A scoping review

**DOI:** 10.1101/2025.02.03.25321593

**Authors:** Christoph Veigl, Benedikt Schnaubelt, Sabine Heider, Andrea Kornfehl, Simon Orlob, Enrico Baldi, Erwin Snijders, Natalie Anderson, Sabine Nabecker, Joachim Schlieber, Zehra’ Al-Hilali, Mahmoud Tageldin Mustafa, Mario Krammel, Federico Semeraro, Robert Greif, Sebastian Schnaubelt

**Affiliations:** Dpt. of Emergency Medicine, Medical University of Vienna, Austria; PULS – Austrian Cardiac Arrest Awareness Association, Austria; Dpt. of Anesthesiology and Intensive Care Medicine, Medical University of Graz, Austria; Institute for Emergency Medicine, University Hospital Schleswig Holstein, Kiel, Germany; Division of Cardiology, Fondazione IRCCS Policlinico San Matteo, Pavia, Italy; Cardiac Arrest and Resuscitation Science Research Team (RESTART), Fondazione IRCCS Policlinico San Matteo, Pavia, Italy; Dpt. of Emergency Medicine, Antwerp University Hospital, Belgium; University of Auckland, New Zealand; Dpt. of Anesthesiology and Pain Management, Mount Sinai Hospital, Toronto, Canada; Dpt. of Anesthesiology and Intensive Care Medicine, AUVA Trauma Center Salzburg, Austria; Arab Resuscitation Council, Dubai, United Arab Emirates; Sudan Resuscitation Council, Khartoum, Sudan; Emergency Medical Service Vienna, Austria; Dpt. of Anesthesia, Intensive Care, and Prehospital Emergency Medicine, Maggiore Hospital Carlo Alberto Pizzardi, Bologna, Italy; Faculty of Medicine, University of Bern, Switzerland; Department of Surgical Science, University of Torino, Torino, Italy

**Keywords:** diversity, obesity, ethnicity, sex, gender, education, training, manikins, cardiac arrest, cardiopulmonary resuscitation, basic life support

## Abstract

Cardiopulmonary resuscitation (CPR) manikins typically appear white, lean, and male. However, internationally, this does not represent the overall population or those who are at greatest risk of cardiac arrest. Diverse demographic groups including people of colour, women, and obese people, are known to be less likely to receive bystander CPR, public access defibrillation, and suffer less favourable outcomes. It is plausible that failure to represent women, racially diverse, and non-lean manikins can contribute to poor clinical outcomes in these populations. The aim of this scoping review was to summarize the current evidence for adaptations of manikins used for layperson Basic Life Support (BLS) training. Data on participant characteristics, manikin adaptations, study design, and key findings of included studies describing or evaluating CPR manikin diversity were extracted. Initially, 2,719 studies were identified and 15 studies were finally included and were grouped into 1) studies analyzing adaptions of “standard” manikins used in training (n=11) and 2) studies evaluating CPR manikin diversity used for online learning and on social media (n=4). Six of the studies analyzing different adaptations reported the influence of the manikins’ sex on comfort in performing CPR, quality of chest compression, AED use, and removing clothes, four the effects of obese manikins, and one an ethnically diverse manikin. Seven of the studies used do-it-yourself adaptions. Racial and gender diversity of CPR manikins found in educational videos was limited, with only 5% of educational videos featuring non-white manikins and 1% featuring female manikins. Adaptations of manikins used for BLS CPR training for laypersons still do not represent the diversity of communities most people are living in, internationally. There are hints that using diverse racial manikins has the potential to improve engagement in CPR training. Reported barriers hindering the use of adapted manikins were high costs and availability of these manikins.

**What is already known on this topic:**

- Manikins for layperson cardiopulmonary resuscitation (CPR) training are typically white, lean, and male all over the world.
- Certain sociodemographic groups experience lower bystander rates, defibrillator use, and lower survival rates after out-of-hospital cardiac arrest.

**What this study adds:**

- The majority of manikins used in studies evaluating adaptions are do-it-yourself adaptions.
- Only one study evaluated the view of ethnically diverse people on racial diverse manikins and how these adaptions should be conducted.

**How this study might affect research, practice or policy:**

- By raising awareness for these sociodemographic disparities in CPR training, educators all over the world should be part of the solution.
- To improve cardiac arrest outcomes among these groups, organisations involved in CPR education should ensure diversity of manikins, and manufacturers should offer low-cost diverse manikins.

## Introduction

Cardiopulmonary resuscitation (CPR) manikins typically appear white, lean and male but internationally, this of course does not represent the overall population or those who are at greatest risk of cardiac arrest. [1] Disparities in both the occurrence of cardiac arrest and the likelihood of resuscitation by witnesses are well known across many characteristics of discrimination. As such, people with different ethnicities, women, and obese people are less likely to receive bystander CPR or public access defibrillation use, resulting in less favourable outcomes. [2–5]

The influence of sex on survival in out-of-hospital cardiac arrest (OHCA) is still under debate even though literature reports a higher survival-to-hospital admission rate in female patients. [6–11] Data on survival-to-hospital discharge and neurological performance are conflicting. [10,12–19] Some studies suggest a better outcome in female patients [6–8,10,11], others show either no survival advantage [9,11,14,18] or higher survival rates in male patients when compared to females. [13,15–17,20–27] A tendency towards unfavourable outcomes for women remains after adjusting for known outcome predictors such as age or initial rhythm, and some authors have even concluded that male sex is independently associated with improved survival. [15,22,23,28,29] The complex underlying factors explaining these differences remain unclear. One explanation could be that women suffering cardiac arrest are significantly less likely to receive potentially lifesaving bystander CPR. [6,12,18,20,30–34] In addition, there is also evidence indicating that a defibrillator is used significantly less frequently in women. [35,36] A recent scoping review on disparities in bystander CPR and education suggested discomfort of laypersons to place their hands on a woman’s chest as a possible reason. [3] Similarly, non-white individuals may be less likely to receive bystander CPR, regardless of their racial or ethnic background or the income level of the neighbourhood where the cardiac arrest occurred. [2,4]

Resuscitation manikins are designed to train CPR skills and offer the opportunity to practice in a controlled, protected environment. However, most manikins used in CPR training are lean, white, and male (or at least male-appearing), and only rarely females, racially diverse, or non-lean. [1,37] This lack of diversity in manikins might affect the application and effectiveness of CPR in real life, and result in a negative impact on clinical outcomes in the respective population subgroups.

The aim of this scoping review was, thus, to identify and describe research evaluating diversified manikins used for layperson BLS education, including self-made adaptations, with the further aim to raise awareness for the problem, provide a basis for stakeholder discussions, and encourage further research and innovation.

## Methods

### Eligibility criteria

This scoping review was performed in accoradance with the Preferred Reporting Items for Systematic Reviews and Meta-analyses Extension for Scoping Reviews (PRISMA-ScR). [38] A specific review protocol including a search strategy was created (PRISMA checklist available as *Supplement S1*).

The PICOST question was:

#### Population

Laypersons or healthcare providers, participating in a BLS training or chest-compressions-only CPR

#### Intervention

BLS training using a diversified manikin (see definition below)

#### Comparison

BLS training using a male, white, lean manikin (referred as “standard” manikin)

#### Outcomes

- Patient / clinical: Return of spontaneous circulation (ROSC), survival to hospital discharge, 30-days survival, 12-months survival, neurological outcome at hospital discharge / 30-days
- Educational: knowledge acquisition, skills acquisition, willingness to perform CPR, participant satisfaction at the end of the respective course and later (3 months, 1 year)

#### Study Design

Included studies: Randomized controlled trials (RCTs) and non-randomized studies (non-randomized controlled trials, controlled before-and-after studies, cohort studies, and case series n ≥ 5), poster presentations, commentaries, editorials. All languages are included as ≥ long as there is an English abstract

Excluded studies: Research on ALS training with high fidelity manikins

#### Timeframe

### Publications from inception to 7^th^ May 2024 with an update on 31^st^ October 2024

This review focused on CPR manikins used in BLS training and not on the training itself. Therefore, we included accredited BLS training (instructor-led courses accredited by resuscitation councils or rescue organisations) as well as other course curricula. We defined “diverse manikins” as all manikins with any changes compared to a male / flat-chested, white, lean manikin. These adaptations might be modified body shapes (e.g., obesity or pregnancy), age (e.g., adult, geriatric, paediatric), modified skin pigment (e.g., ethnic differences), or gendering the manikin (e.g., long hair, make-up, breast tissue to simulate female body features).

### Information sources and search strategy

The search was conducted on 7^th^ May 2024, with an update on 31^st^ October 2024. The search strategy was developed by two information specialists (CR and BH, Medical University of Vienna, Austria)—see *Supplement S2*. Records from database searches were downloaded and imported in Rayyan (https://www.rayyan.ai/) to facilitate the removal of duplicates and screening. Databases searched included MEDLINE (R) ALL (Ovid), Embase (Ovid), PsycINFO (Ovid), CINAHL (Ebscohost), ERIC (Ebscohost), Web of Science (Clarivate), Infromit, Scopus (Elsevier); and Cochrane Central Register of Controlled Trials (Cochrane Library via Wiley Online). Hand-searching included reference lists of included studies and a keyword search of the social media platform X.

### Study selection process

After de-duplication, titles and abstracts were independently screened by two reviewers (CV, SS). Conflicts were resolved via discussion and agreement between the two reviewers. The process was followed by a full-text assessment of the potentially relevant papers by the same two reviewers.

### Data charting process, data items, and synthesis of results

Information from selected articles was extracted into a data extraction sheet which included details of the first author, country, publication year, article type, study design, main topics and/or manikin adaptations, participants, and the key findings of the studies. Data extraction was reviewed by the co-authors, and disagreements were discussed until consensus was reached.

## Results

### Study screening and selection

The search found 2,719 records, and one additional record was identified through X (former Twitter). After deleting 1,444 duplicates, the titles and abstracts of the remaining 1,275 records were screened, and 21 articles were entered into full-text assessment. Finally, 15 articles were included for data extraction and analysis (*Figure 1* PRISMA-ScR flow diagram).

**Figure 1:**
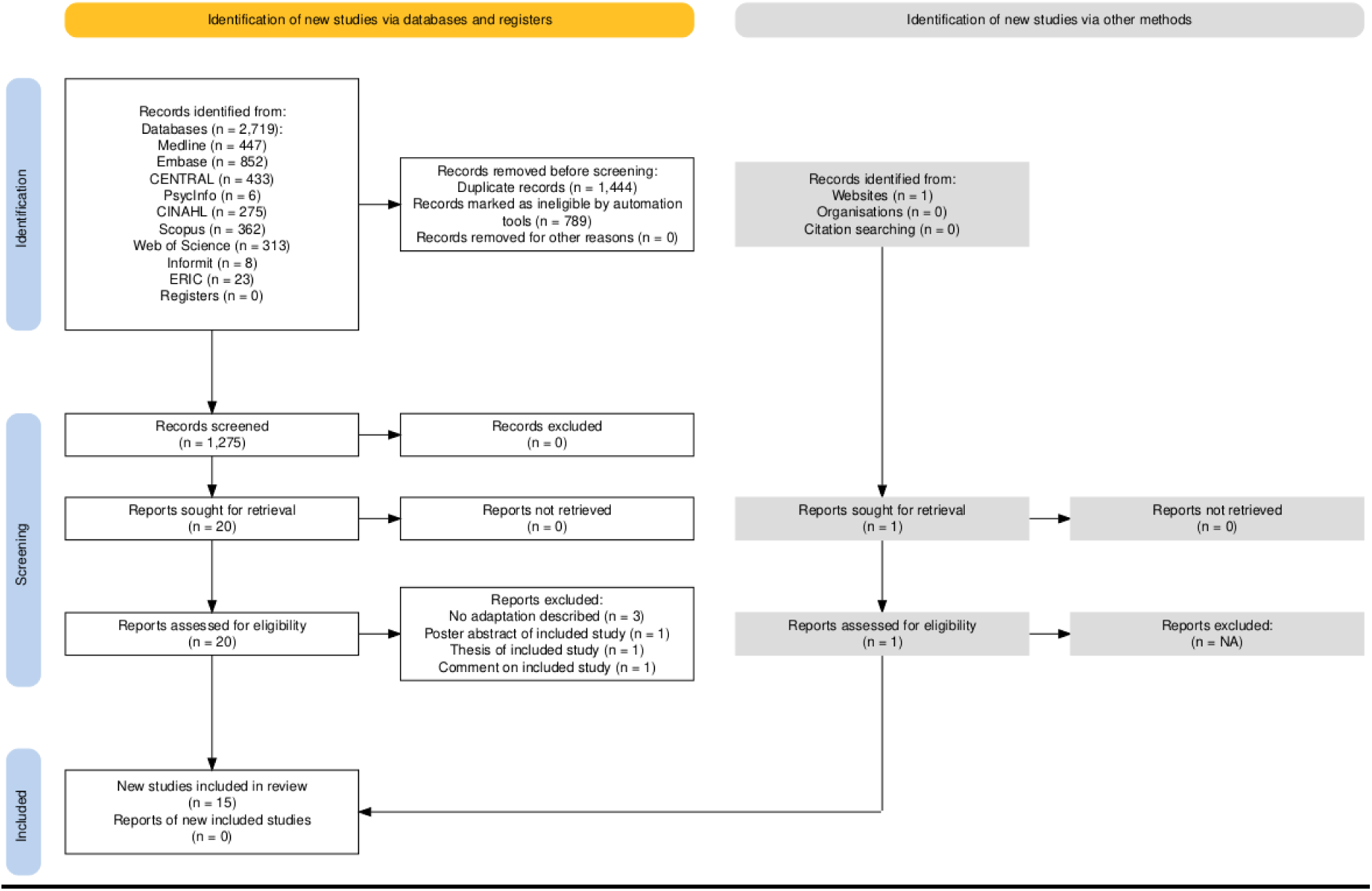
PRISMA flow diagram created with https://estech.shinyapps.io/prisma_flowdiagram/ [60].

### Study characteristics

The geographical distribution and respective income classifications as per definition of the World Bank are summarized in *Table 1*. [39] All included publications described empirical research. All studies were conducted in high-income countries, 60% were published in the last three years (n= 9) and the first studies (n= 2) were published in 2014. Ten publications were original research articles, [1,40–48] three were published as a conference abstract, [49–51] one as a conference poster [52] and one as a research letter [37].

**Table 1:** Included studies per geographical region in alphabetical order.

|  |  |  |
| --- | --- | --- |
| **Region** | **No. of studies** | **Countries** |
| East Asia and Pacific | 3 | Australia (2), Japan (1) - *all high income*<sup>1</sup> |
| Europe | 3 | Germany (1), Ireland (1), UK (1) – *all high income*<sup>1</sup> |
| North America | 9 | Canada (3), USA (6) – *all high income*<sup>1</sup> |
| **Total** | **15** |  |

Two groups of publications according to the study focus were formed: One analyzing effects of diverse manikin use in in-person BLS training including chest-compression-only CPR (11 included studies) (*Table 2*) [40–46,49–52]; and the other focusing on representation of diverse CPR manikins for layperson education on social media (four included studies) (*Table 3)* [1,37,47,48]. An overview of the most important findings can be found in *Figure 2*.

**Figure 2:**
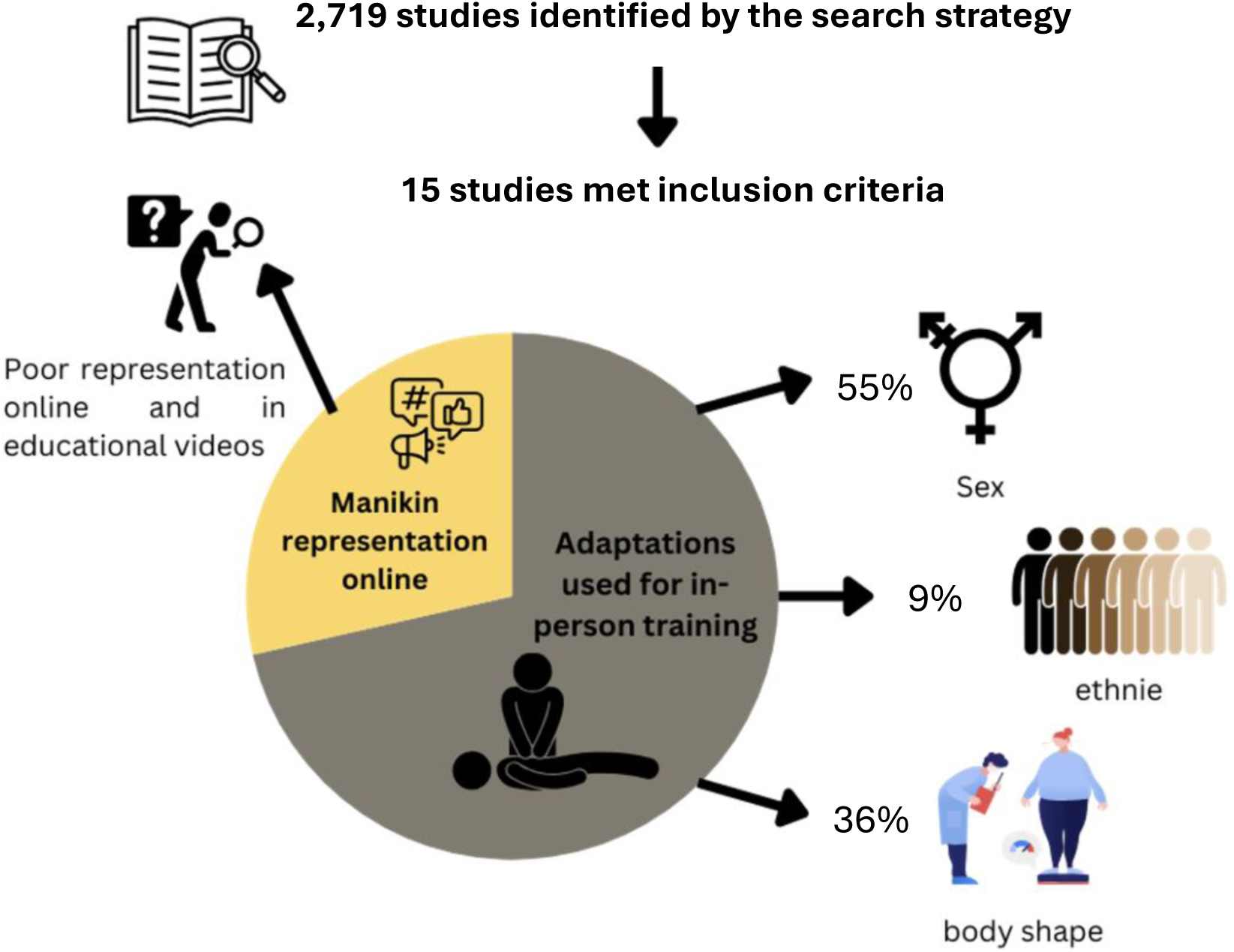
Overview of the results of the included studies.

**Table 2:** Included publications analyzing different adaptations of manikins for in-person BLS training.

BLS= Basic Life Support; CPR= cardiopulmonary resuscitation; DIY= do-it-yourself; AED= automated external defibrillator; BMI= body mass index.
|  |  |  |  |  |  |
| --- | --- | --- | --- | --- | --- |
| **First author, Publication year, Country** | **Article and Research Typ** | **Main Topic** | **Methods / Participants** | **Manikin adaptation** | **Relevant key findings** |
| K. Goulding, 2022, Australia [42] | Original research article<br>Prospective blinded randomised-controlled crossover study | Improvement of chest compressions using a BariBoard™ (Iron Duck®, Chicopee, USA) on a modified manikin simulating a morbidly obese patient (BMI = 40 kg/m2). | Medical professionals from the critical care department.<br>201 participants | DIY modifications of manikin by putting vacuum sealed bags of porcine fat were inserted into anterior and posterior pockets of a customised neoprene suit, which was placed on a resuscitation model to emulate a BMI of 40 kg/m2. | Significant higher overall efficacy of compressions in obese manikin and for adequate compression depth and recoil.<br>No statistically significant difference between the proportions of participants achieving an adequate compression rate.<br>After adjustment for confounders and interactions, there was no difference in overall efficacy between both groups. |
| M. Kraft, 2024, Germany [51] | Conference abstract<br>Prospective randomized-controlled cross-over study | Influence of manikin sex on chest compression rate, depth, recoil, and hand-off time. | Layperson without medical background performed chest compression-only CPR for eight minutes<br>198 participants were randomized 1:1 either to male or female manikin first | No further information about adaptations given | Significant higher average compression rate in females vs. male manikins (104 (±19) bpm vs 97 (±28) bpm, p=0.001).<br>Significant lower number of insufficient recoil in female vs. male manikins (13% (±22%) vs. 24% (±29%), p<0.001).<br>No statistically significant differences in the remaining CPR metrics. |
| C. E. Kramer, 2014, Canada [41] | Original research article<br>Randomized controlled trial | • Influence of manikin sex on removing clothes and placement of rescuer hands of layperson. | • Undergraduate students<br>• Participants randomly assigned to do CPR with an AED on female or male manikin.<br>• 69 participants: 36 trained with standard and 33 with female manikin | • DIY modifications of manikin using silicone breasts, a wig, make-up, a front-opening brassiere and colour-coordinated women's clothing. | • Significantly more participants on the male manikin removed clothes completely (92% vs. 42%).<br>• Female participants assigned to female manikin removed significantly more often all clothes than male participants (67% vs. 13%). No difference in removing clothes between men and women in male manikin.<br>• Hand placement on male manikin showed more variability over a greater area of the chest, while hand placement on female manikin tended to be centred between the breasts. |
| P. J. Secombe, 2018, Australia [44] | Original research article<br>Prospective randomised controlled crossover study | • Adequacy of thoracic compressions on a modified manikin simulating a morbidly obese patient (BMI = 40 kg/m<sup>2</sup>). compared to a "standard" non-obese manikin and participant-perceived measures of effectiveness, fatigue and pain | • Medical professionals from the critical care department at Alice Springs Hospital, Australia.<br>• 101 participants | • DIY modifications of "standard" manikin by putting vacuum sealed bags of porcine fat were inserted into anterior and posterior pockets of a customised neoprene suit, which was placed on a resuscitation model to emulate a BMI of 40 kg/m<sup>2</sup>. | • Significant lower adequacy for chest compression on obese manikin, and for each component of primary outcome as well.<br>• No significant difference in the mean chest compression rate, but highly significant difference in both the mean compression depth achieved overall and, in each quartile, and the mean release velocity.<br>• There was a significant difference in perceived effectiveness, fatigue and pain favouring the non-obese manikin. |
| A. Tellson, 2017, USA [45] | Original research article<br>Randomized controlled trial | • Evaluate chest compression for three adult simulation manikins (“standard”, obese, and morbidly obese)<br>• Contribution of participant characteristics of height, weight, sex, and upper body strength to the quality of chest compressions in obese and morbidly obese manikins. | • Medical professionals from a for-profit cardiovascular specialty hospital<br>• 61 participants<br>• The thorax framework and the sensors from a standard CPR manikin were in the correct anatomical position in the adapted manikins. | • Manikins to mimic an obese and a morbidly obese adult were created, manufactured, and tested by a professional in the field of manikin design and manufacturing by adding the calculated amount of a foam substance of a consistency similar to adipose tissue. | • Significant more successful chest compressions on “standard” manikin, compared to both obese and morbidly obese manikins.<br>• No significant difference in performance between the two obese manikins.<br>• BMI of rescuer, upper body strength, ethnicity, and manikin order were all significantly associated with the number of successful compressions. |
| R. T. Kim, 2023, USA [46] | Original research article<br>Non-randomized controlled trial (5 of 31 manikins were adapted) | • To evaluate the influence of the use of a female manikin during CPR training on reported comfort levels pre- and post-training of trainees in performing CPR on women with a questionnaire. | • Layperson as part of an annual training<br>• 79 participants: 58 using standard manikin and 21 female manikin<br>• Pre-training and post-training survey | • DIY modifications of “standard” manikin with breast adjunct constructed from a lifecast silicone mold. | • In the post-training survey comparison, trainees in the adjunct group reported greater comfort in performing CPR on women, compared to the control group.<br>• This effect persisted for questions relating to both clothed and unclothed women. |
| K. Kobayashi, 2024, Japan [43] | Original research article<br>Cross-over study | • Effect of manikin sex on the quality of chest compressions. | • Students studying medical care and welfare without prior training.<br>• 34 participants (15 males, 19 females) | • DIY modifications of “standard” manikin by using silicone breasts and dressed in a brassiere, long skirt, long-sleeved shirt, and long-haired wig. | • Significant higher appropriate compression depth ratio (%) and significant lower appropriate recoil ratio (%) of chest compressions on female manikins.<br>• No significant difference in other outcomes. |
| J. A. Longo, 2024, USA [40] | Original research article<br>Cross-over study | • Ability of to perform chest compressions on an obese manikin.<br>• Analyze the effect of American football protective equipment on the performance of chest compressions by emergency responders | • Emergency responders performing 2 minutes of chest compressions on a “standard” and an obese manikin, once with and once without chest/shoulder pads on.<br>• 50 participants | • Obese patient was simulated using a Nasco Bariatric CPR Manikin (Nasco Healthcare Inc. Life/form®, Saugerties, USA) (commercially available). | • Mean compression depth significantly lower when performed on the obese manikin compared to “standard” manikin, while chest recoil was significantly better in obese manikin.<br>• No significant difference in chest compression rate. |
| M. Leary, 2018, USA [49] | Conference abstract<br>Randomized controlled trial | • Evaluate laypersons response to an unannounced simulated virtual reality cardiac arrest based on the victim’s sex. | • Untrained laypersons using a virtual reality cardiac arrest system to simulate victim's sex (female or male), while performing CPR on the “standard” manikin.<br>• 75 participants | • DIY immersive virtual reality system to simulate the sex of virtual manikin. | • Non significant difference between CPR performed on male vs on female victims (65% vs. 54%).<br>• Non significant difference between AED use for male vs female victims (21% vs. 15%). |
| P. O'Hare, 2014, Ireland [50] | Conference abstract<br>Randomized controlled trial | • Effect of patient sex to time-to-first shock by untrained bystanders using a public access defibrillator. | • Untrained laypersons<br>• 78 participants<br>• Half of the participants trained with a "female" manikin clothed in a front-opening hooded sweater with a bra underneath and another half with standard manikin. | • DIY modifications of “standard” manikin with a bra under sweater. | • All participants successfully delivered a shock.<br>• No statistically significant difference between the times to first shock of the female and male clothed manikin groups.<br>• 88.5% of the participants were able to correctly place the electrodes. |
| J. Walls, 2023, United Kingdom [52] | Conference poster<br>Observational, questionnaire study | • Observe choice of manikin between the “standard” manikin or black OBI manikin within ethnic groups and receive feedback on their choice.<br>• Record opinions regarding the need for ethnically diverse manikins for BLS Training. | • Healthcare Professionals<br>• 30 participants | • Brayden OBI manikin (Innosonian Europe, Farnborough, United Kingdom) (commercially available) representing an ethnically appropriate adult black male with anatomically correct features. | • From White or Black backgrounds 56% chose black manikin, 60% from Asian group chose this manikin, and only 25% participant from the Mixed group (White Asian or Other) chose the diverse manikin.<br>• 56% of Black participants stated that they identified more with black manikin, 33% said neither and 11% said "standard" manikin.<br>• 78% of Black participants thought it important that manikins reflect ethnicity. The same number felt that having black manikins will help improve CPR.<br>• Most participants identifying as ‘black,’ strongly agreed that ethnically accurate manikins will help to improve CPR training. |

**Table 3:**
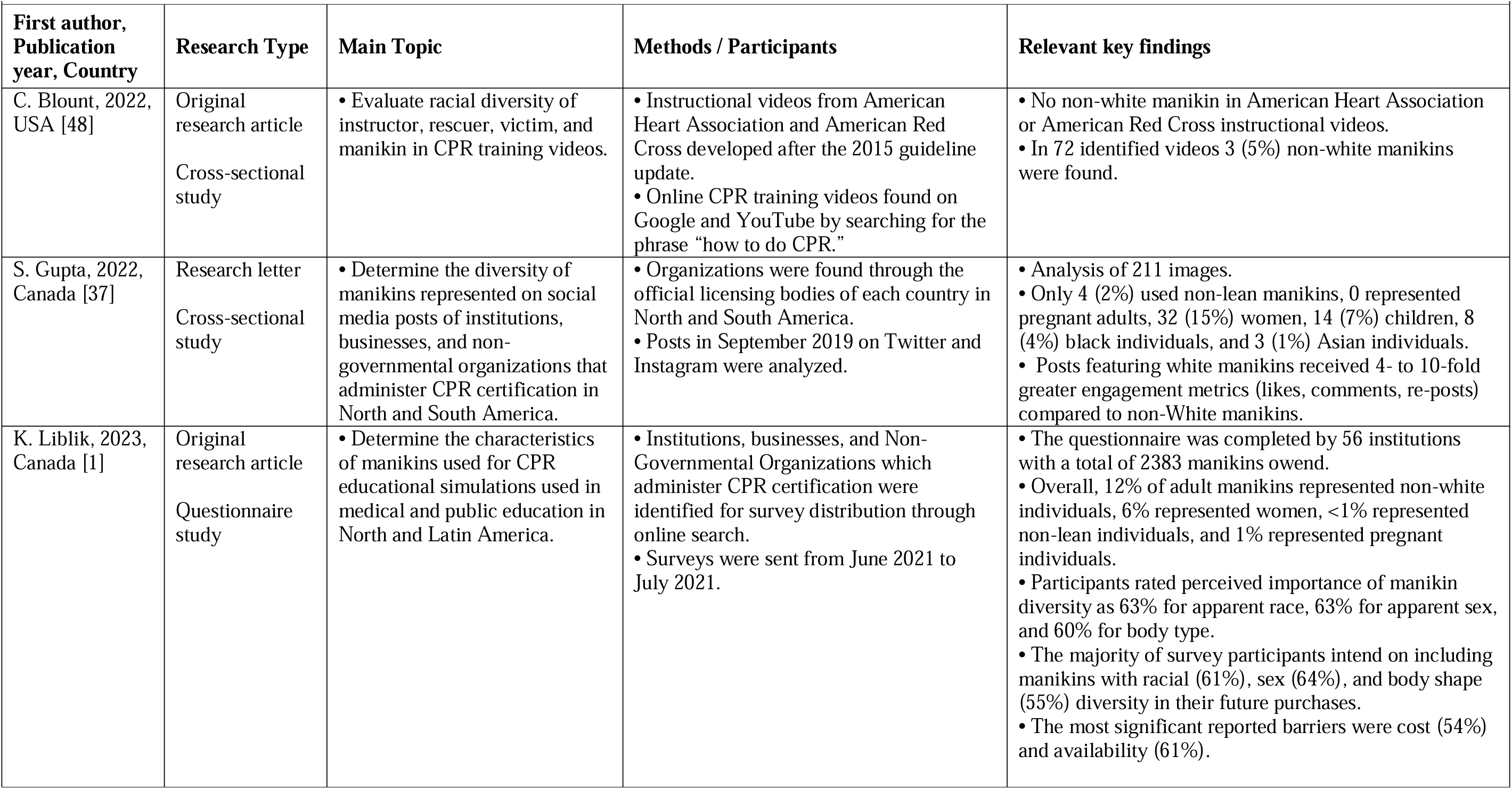
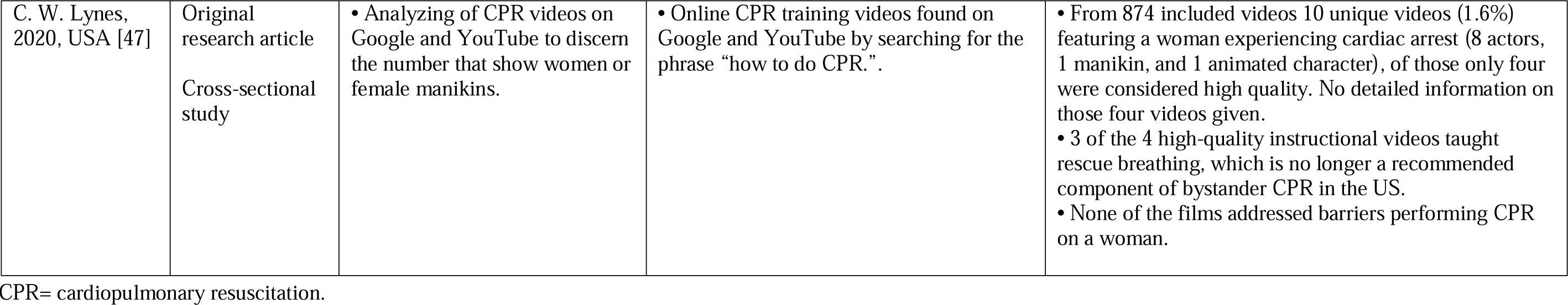
Included publications analyzing the diversity of CPR manikins used in educational videos and social media.

| **First author, Publication year, Country** | **Research Type** | **Main Topic** | **Methods / Participants** | **Relevant key findings** |
| --- | --- | --- | --- | --- |
| C. Blount, 2022, USA [48] | Original research article<br>Cross-sectional study | • Evaluate racial diversity of instructor, rescuer, victim, and manikin in CPR training videos. | • Instructional videos from American Heart Association and American Red Cross developed after the 2015 guideline update.<br>• Online CPR training videos found on Google and YouTube by searching for the phrase “how to do CPR.” | • No non-white manikin in American Heart Association or American Red Cross instructional videos.<br>• In 72 identified videos 3 (5%) non-white manikins were found. |
| S. Gupta, 2022, Canada [37] | Research letter<br>Cross-sectional study | • Determine the diversity of manikins represented on social media posts of institutions, businesses, and non-governmental organizations that administer CPR certification in North and South America. | • Organizations were found through the official licensing bodies of each country in North and South America.<br>• Posts in September 2019 on Twitter and Instagram were analyzed. | • Analysis of 211 images.<br>• Only 4 (2%) used non-lean manikins, 0 represented pregnant adults, 32 (15%) women, 14 (7%) children, 8 (4%) black individuals, and 3 (1%) Asian individuals.<br>• Posts featuring white manikins received 4- to 10-fold greater engagement metrics (likes, comments, re-posts) compared to non-White manikins. |
| K. Liblik, 2023, Canada [1] | Original research article<br>Questionnaire study | • Determine the characteristics of manikins used for CPR educational simulations used in medical and public education in North and Latin America. | • Institutions, businesses, and Non-Governmental Organizations which administer CPR certification were identified for survey distribution through online search.<br>• Surveys were sent from June 2021 to July 2021. | • The questionnaire was completed by 56 institutions with a total of 2383 manikins owned.<br>• Overall, 12% of adult manikins represented non-white individuals, 6% represented women, <1% represented non-lean individuals, and 1% represented pregnant individuals.<br>• Participants rated perceived importance of manikin diversity as 63% for apparent race, 63% for apparent sex, and 60% for body type.<br>• The majority of survey participants intend on including manikins with racial (61%), sex (64%), and body shape (55%) diversity in their future purchases.<br>• The most significant reported barriers were cost (54%) and availability (61%). |
| C. W. Lynes,<br>2020, USA [47] | Original<br>research article<br>Cross-sectional<br>study | Analyzing of CPR videos on Google and YouTube to discern the number that show women or female manikins. | Online CPR training videos found on Google and YouTube by searching for the phrase “how to do CPR.”. | From 874 included videos 10 unique videos (1.6%) featuring a woman experiencing cardiac arrest (8 actors, 1 manikin, and 1 animated character), of those only four were considered high quality. No detailed information on those four videos given.<br>3 of the 4 high-quality instructional videos taught rescue breathing, which is no longer a recommended component of bystander CPR in the US.<br>None of the films addressed barriers performing CPR on a woman. |

### Effects of manikin diversity

From the eleven studies evaluating different manikin adaptions (totally including 976 participants [40–46,49–52]), six (55%) investigated the influence of manikin sex, [41,43,46,49–51], four (36%) evaluated the effects of an obese manikin, [40,42,44,45] and one (9%) focused on ethnically diverse manikins. [52] All of the female or obese manikins represented a white person. Seven studies (64%) adapted a “standard” manikin to produce a diverse manikin (do-it-yourself, DIY-manikin) [41–44,46,49,50], two studies used commercially available manikins: one an obese patient [40] and the other an ethnically appropriate adult black male [52], but none represented a female patient. One study used a manikin produced by a professional manikin manufacturer, but not commercially available at the time of publication [45], and another study did not specify how they carried out the adjustment. [51] All other included studies focusing on the effect of the manikins’ sex reported a DIY adaptation of a “standard” manikin. [41,43,46,50] There was a great variability of adaptations to represent a female manikin, ranging from the use of silicone breasts [41,43,46], a wig, make-up, a front-opening brassiere and clothing perceived as “typically” female, [41,43] to just small modifications such as a bra under a sweater. [50]

Five of the studies (45%) observed training for healthcare professionals. [40,42,44,45,52] Four of these assessed chest compression quality. [40,42,44,45] One observed participants’ choice between a standard or a racially diverse manikin for training, and found that for the 78% of participants who identified as black, it was important that they were represented in manikins and that reflecting ethnicity would improve engagement in CPR training. [52]

All studies assessing laypersons as participants focused on the influence of sex and reported a variety of outcomes: the use of an AED and the time to first shock [49,50], bystander CPR initiation [49], removing clothes and hand placement during chest compressions [41], chest compression quality [43,51], and the effect of female manikins on comfort in performing CPR [46]. In one study, the sex of the patient was simulated using virtual reality, while the participants were performing CPR on a “standard” manikin. [49] The key findings of all the studies are summarized in *Table 2*.

### Presence of manikin diversity in educational videos and on social media

Four publications were included in this group. [1,37,47,48] Three were cross-sectional studies. [37,47,48] One of them [37] determined the diversity of manikins represented in social media posts of institutions and organizations, the other two evaluated diversity regarding race [48] and sex [47] of characters in CPR training videos. The two studies focusing on CPR training videos included official training videos of the American Heart Association (AHA) and American Red Cross (ARC), as well as videos found on YouTube and Google. [47,48] Both studies together included 960 educational CPR videos. One evaluated 86 videos for racial diversity [48] and found three videos (5% of all manikins) using non-white manikins, but no racially diverse manikins in AHA or ARC instructional videos. Another study screened 874 videos for female manikins and found ten unique videos (1%) with female manikins. Four of them (0.5% of all videos) were considered high-quality. [47]

In one study, authors conducted a survey in 56 institutions in North and Latin America to determine the characteristics of manikins used for CPR education. [1] They found 12% of adult manikins represented diverse manikins in participating institutions and identified high costs and availability of diverse manikins as the main barriers to use diverse manikins.

*Table 3* summarizes the key findings.

## Discussion

This scoping review identified 14 publications addressing CPR manikin diversity for BLS training, published between 2014 and 2024. Due to the heterogeneity of the covered topics, we did not conduct a systematic review. Issues addressed were the quality of chest compressions in obese manikins, assessing laypersons’ responses to a cardiac arrest based on the victim’s sex including removing clothes, AED use, and hand placement, and the participants’ choices of manikin between a standard manikin or a dark-skinned manikin.

Bystander CPR rate and survival rates after cardiac arrest in black communities have in the past been reported to be lower than in other communities. [2] However, only four studies included in this review (three were observational [1,37,48]) reported on racial diversity of CPR manikins [1,37,48,52]. Only one of them evaluated the use of non-white manikins (an adult black male with anatomically adapted features). [52]

Most currently commercially available dark-skinned manikins do not accurately reflect the differences between Caucasian and other ethnicities, as they are simply differently skin-coloured manikins. Specific ethnically diverse manikins developed together with respective population groups could possibly improve further tailored resuscitation education, with the potential to increase engagement and willingness to help in these groups. [52–54] For instance, CPR teaching participants self-identifying as black stated that manikin reflecting ethnicity would improve engagement in CPR training. [52]

Suprisingly only two studies used commercially available manikins [40,52], and none of these were female. Reason behind that could be non-availability with manufacturers or too high costs for CPR mass education. This assumption is reinforced by a survey conducted among organisations teaching CPR, in which the most significant reported barriers were cost and availability of diverse manikins. [1] A novel way to overcome the lack of availability could be the use of new technologies like augmented or virtual reality. A recently published study tested a virtual reality system simulating a female patient and providing visual and physical feedback. [55] Although these technologies have already found their way into many medical fields including CPR education [49,56–58], there are still significant barriers such as required resources and implementation before they can be broadly available.

Sex-diverse CPR manikins could familiarize rescuers with CPR on women and might reduce barriers towards chest compressions, which could finally increase the possibility of survival for women in cardiac arrest. [46]. Furthermore, concerns about inappropriate touching women’s breasts, accusations of sexual assault, and fear of causing injury by starting bystander CPR in women [31] could be specifically addressed during the CPR training.

Until affordable diverse CPR manikins are available, CPR training organizations might create their own diverse DIY manikins. An online tutorial estimates the costs to adapt a “standard” manikin to a female manikin to be about 6 US dollars, [59] and a a recently published scoping review provides an overview of DIY-devices for layperson CPR training. [60]

### Gaps of Knowledge and Action Points

The educational impact on patient survival using different diverse manikins during CPR training is unknown. The degree of fidelity and to which detail manikins need to be adapted are unclear. Properly powered high-stake studies to answer these two knowledge gaps are certainly needed.

Also, CPR training organizations and instructors need to raise awareness for sociodemographic disparities in CPR outcomes. The use of “standard” manikins for CPR training stands in a wide context of implicit bias, which is unconscious and unintentional mental associations that impact our understanding and actions. Implicit bias is often difficult to recognise and to address during teaching. To reduce inequities in training, there are some evidence-informed teaching tips and strategies regarding recognition of implicit bias and management for medical educators described in the literature. [61] As resuscitation education becomes more sophisticated, the idea of a white, male, lean manikin as the only manikin used should be abandoned in favour of manikins reflecting the diversity of the communities. All stakeholders involved in resuscitation education should contribute to the solution.

## Limitations

Four of the included studies were only available as an abstract. Also, only a limited number of studies report on diversity of manikins during CPR training and no found publication reflected the view from low resource settings. However, in such settings other health care problems that need to be addressed might have a higher priority. Moreover, there may be real-life CPR situations in which a gap between biological sex and the sex perceived by the attending layperson exists – this could be addressed in further research. The authors’ list of this scoping review is not as diverse as potentially possible, which might have conveyed biased views.

## Conclusion

This scoping review provides limited evidence on the effect of the use of various diverse manikins (e.g., concerning race, sex, age, body shape, etc.) for CPR education. The often used white male manikins do not represent the diversity of communities trained in CPR or suffering cardiac arrest. The vast majority of found studies reported on adaptations of manikins in a do-it-yourself way to generate different diverse manikins. The most prominent reported barrier not to use diverse manikins were high costs and availability of such manikins.

Using diverse racial manikins has the potential to improve engagement in CPR training in population subgroups with the aim to increase survival of cardiac arrest. Such manikins can raise awareness of sociodemographic disparities in CPR outcome during training and reduce barriers not to help.

## Supporting information

Supplemental 1 - PRISMA Checklist

## Data Availability

All data produced in the present study are available upon reasonable request to the authors

## Acknowledgements

The authors acknowledge the assistance provided by the information specialists of the Medical University of Vienna Birgit Heller and Caroline Reitbrecht for building the searching strategy.

## Patient and Public Involvement statement

Patient and public involvement in this review was not feasible.

## Ethics statements

### Ethics approval and patient consent for publication

For this scoping review no personal, sensitive or confdential information from participants was collected and only publicly accessible documents were used as evidence. Therefore, no ethics approvel or patient consents for publication are needed.

