## Supplemental Data 1 for "Diversity of CPR manikins for basic life support education: Use of manikin sex, race, and body shape – A scoping review"

**Supplement S2 – Search results and strategies**

| **Database** | **Results** |
| --- | --- |
| Medline | 447 |
| Embase | 852 |
| CENTRAL | 433 |
| PsycInfo | 6 |
| CINAHL | 275 |
| Scopus | 362 |
| Web of Science | 313 |
| Informit | 8 |
| ERIC | 23 |
| **Total (incl. dublicates)** | **2719** |

**Search History Medline conducted at 7th May 2024**

| **#** | **Query** | **Results** |
| --- | --- | --- |
| 1 | Cardiopulmonary Resuscitation/ | 22390 |
| 2 | defibrillators/ | 2279 |
| 3 | ("Cardiopulmonary Resuscitation*" or "Cardio pulmonary Resuscitation*" or "cardiopulmonary reanimation*" or "cardio pulmonary reanimation*" or CPR or "basic life support*" or "cardiac life support*" or BLS or "chest compression*" or "cardiac massage*" or "heart massage*" or "mouth to mouth Resuscitation*" or defibrillator* or AED).ti,ab,kf. | 65711 |
| 4 | 1 or 2 or 3 | 73794 |
| 5 | exp health inequities/ | 41376 |
| 6 | exp cultural diversity/ | 13465 |
| 7 | exp sex/ | 7759 |
| 8 | exp ethnicity/ | 110672 |
| 9 | exp racial groups/ | 107257 |
| 10 | exp race factors/ | 1315 |
| 11 | exp infant/ | 1273791 |
| 12 | exp child/ | 2203033 |
| 13 | exp aged/ | 3500238 |
| 14 | somatotypes/ | 2323 |
| 15 | Sociodemographic Factors/ | 716 |
| 16 | 5 or 6 or 7 or 8 or 9 or 10 or 11 or 12 or 13 or 14 or 15 | 6217050 |
| 17 | manikins/ | 5738 |
| 18 | 16 and 17 | 976 |
| 19 | ((manikin* or mannikin* or mannequin* or dummy or dummies or "simulated patient*" or "patient simulator*" or "CPR doll*" or PractiMan or Prestan or LifeSaveHer or Simulaids or "Life form" or "Nasco Healthcare" or Laerdal or WorldPoint or Simulaids or womanikin or "Resusci Anne") adj10 (diversity or diverse or variation* or represent* or sex or gender* or woman or women or female* or breast* or race or racial or ethnic* or minorit* or black* or asian* or hispanic* or "non white*" or "people of color" or "people of colour" or "person of color" or "person of colour" or POC or WOC or BIPOC or "skin color*" or "skin colour*" or age or infant* or newborn* or neonat* or baby or babies or child* or pediatr* or paediatr* or aged or elder* or geriatric* or "old person*" or "old people" or appearance* or "body shape*" or "body habit*" or "body build*" or "body type*" or somatotype* or obese or obesity or overweight* or bariatric* or "non lean" or pregnan* or socioeconomic* or sociodemographic* or demographic* or bias*)).ti,ab,kf. | 2131 |
| 20 | 18 or 19 | 2736 |
| 21 | 4 and 20 | **439** |

**Search History Embase conducted at 7th May 2024**

| **No.** | **Query** | **Results** |
| --- | --- | --- |
| #22 | #4 AND #21 | **828** |
| #21 | #19 OR #20 | 3519 |
| #20 | ((manikin* OR mannikin* OR mannequin* OR dummy OR dummies OR 'simulated patient*' OR 'patient simulator*' OR 'cpr doll*' OR practiman OR prestan OR lifesaveher OR simulaids OR 'life form' OR 'nasco healthcare' OR laerdal OR worldpoint OR simulaids OR womanikin OR 'resusci anne') NEAR/10 (diversity OR diverse OR variation* OR represent* OR sex OR gender* OR woman OR women OR female* OR breast* OR race OR racial OR ethnic* OR minorit* OR black* OR asian* OR hispanic* OR 'non white*' OR 'people of color' OR 'people of colour' OR 'person of color' OR 'person of colour' OR poc OR woc OR bipoc OR 'skin color*' OR 'skin colour*' OR age OR infant* OR newborn* OR neonat* OR baby OR babies OR child* OR pediatr* OR paediatr* OR aged OR elder* OR geriatric* OR 'old person*' OR 'old people' OR appearance* OR 'body shape*' OR 'body habit*' OR 'body build*' OR 'body type*' OR somatotype* OR obese OR obesity OR overweight* OR bariatric* OR 'non lean' OR pregnan* OR socioeconomic* OR sociodemographic* OR demographic* OR bias*)):ti,ab,kw | 2983 |
| #19 | #15 AND #18 | 842 |
| #18 | #16 OR #17 | 4779 |
| #17 | 'patient simulator'/exp | 1170 |
| #16 | 'manikin'/de | 3674 |
| #15 | #5 OR #6 OR #7 OR #8 OR #9 OR #10 OR #11 OR #12 OR #13 OR #14 | 8514691 |
| #14 | 'sociodemographics'/de | 22280 |
| #13 | 'body build'/de | 6409 |
| #12 | 'aged'/exp | 3951582 |
| #11 | 'child'/exp | 3480402 |
| #10 | 'infant'/exp | 1323179 |
| #9 | 'ethnic or racial aspects'/exp | 361333 |
| #8 | 'ancestry group'/exp | 456167 |
| #7 | 'gender and sex'/exp | 1299602 |
| #6 | 'cultural diversity'/exp | 4577 |
| #5 | 'health disparity'/exp | 39731 |
| #4 | #1 OR #2 OR #3 | 209875 |
| #3 | 'cardiopulmonary resuscitation*':ti,ab,kw OR 'cardio pulmonary resuscitation*':ti,ab,kw OR 'cardiopulmonary reanimation*':ti,ab,kw OR 'cardio pulmonary reanimation*':ti,ab,kw OR cpr:ti,ab,kw OR 'basic life support*':ti,ab,kw OR 'cardiac life support*':ti,ab,kw OR bls:ti,ab,kw OR 'chest compression*':ti,ab,kw OR 'cardiac massage*':ti,ab,kw OR 'heart massage*':ti,ab,kw OR 'mouth to mouth resuscitation*':ti,ab,kw OR defibrillator*:ti,ab,kw OR aed:ti,ab,kw | 105269 |
| #2 | 'automated external defibrillator'/de | 3149 |
| #1 | 'resuscitation'/exp | 142789 |

**Search History CENTRAL conducted at 31^st^ October 2024**

| 1 | Cardiopulmonary Resuscitation/ | 1691 |
| --- | --- | --- |
| 2 | defibrillators/ | 130 |
| 3 | ("Cardiopulmonary Resuscitation*" or "Cardio pulmonary Resuscitation*" or "cardiopulmonary reanimation*" or "cardio pulmonary reanimation*" or CPR or "basic life support*" or "cardiac life support*" or BLS or "chest compression*" or "cardiac massage*" or "heart massage*" or "mouth to mouth Resuscitation*" or defibrillator* or AED).ti,ab,kw. | 9174 |
| 4 | 1 or 2 or 3 | 9514 |
| 5 | exp health inequities/ | 296 |
| 6 | exp cultural diversity/ | 109 |
| 7 | exp sex/ | 43 |
| 8 | exp ethnicity/ | 3496 |
| 9 | exp racial groups/ | 1339 |
| 10 | exp race factors/ | 50 |
| 11 | exp infant/ | 45653 |
| 12 | exp child/ | 81664 |
| 13 | exp aged/ | 279840 |
| 14 | somatotypes/ | 24 |
| 15 | Sociodemographic Factors/ | 17 |
| 16 | 5 or 6 or 7 or 8 or 9 or 10 or 11 or 12 or 13 or 14 or 15 | 383558 |
| 17 | manikins/ | 1239 |
| 18 | 16 and 17 | 254 |
| 19 | ((manikin* or mannikin* or mannequin* or dummy or dummies or "simulated patient*" or "patient simulator*" or "CPR doll*" or PractiMan or Prestan or LifeSaveHer or Simulaids or "Life form" or "Nasco Healthcare" or Laerdal or WorldPoint or Simulaids or womanikin or "Resusci Anne") adj10 (diversity or diverse or variation* or represent* or sex or gender* or woman or women or female* or breast* or race or racial or ethnic* or minorit* or black* or asian* or hispanic* or "non white*" or "people of color" or "people of colour" or "person of color" or "person of colour" or POC or WOC or BIPOC or "skin color*" or "skin colour*" or age or infant* or newborn* or neonat* or baby or babies or child* or pediatr* or paediatr* or aged or elder* or geriatric* or "old person*" or "old people" or appearance* or "body shape*" or "body habit*" or "body build*" or "body type*" or somatotype* or obese or obesity or overweight* or bariatric* or "non lean" or pregnan* or socioeconomic* or sociodemographic* or demographic* or bias*)).ti,ab,kw. | 1268 |
| 20 | 18 or 19 | 1367 |
| 21 | 4 and 20 | 433 |

**Search History PsycInfo conducted at 7th May 2024**

| **#** | **Query** | **Results** |
| --- | --- | --- |
| 1 | Cardiopulmonary Resuscitation.mh. | 300 |
| 2 | defibrillators.mh. | 16 |
| 3 | ("Cardiopulmonary Resuscitation*" or "Cardio pulmonary Resuscitation*" or "cardiopulmonary reanimation*" or "cardio pulmonary reanimation*" or CPR or "basic life support*" or "cardiac life support*" or BLS or "chest compression*" or "cardiac massage*" or "heart massage*" or "mouth to mouth Resuscitation*" or defibrillator* or AED).ti,ab,hw,id. | 3629 |
| 4 | 1 or 2 or 3 | 3693 |
| 5 | health inequities.mh. | 24 |
| 6 | cultural diversity.mh. | 2970 |
| 7 | sex.mh. | 335 |
| 8 | ethnicity.mh. | 887 |
| 9 | racial groups.mh. | 235 |
| 10 | race factors.mh. | 93 |
| 11 | infant.mh. | 31999 |
| 12 | child.mh. | 150882 |
| 13 | aged.mh. | 217334 |
| 14 | somatotypes.mh. | 171 |
| 15 | Sociodemographic Factors.mh. | 32 |
| 16 | 5 or 6 or 7 or 8 or 9 or 10 or 11 or 12 or 13 or 14 or 15 | 381479 |
| 17 | manikins.mh. | 175 |
| 18 | 16 and 17 | 27 |
| 19 | ((manikin* or mannikin* or mannequin* or dummy or dummies or "simulated patient*" or "patient simulator*" or "CPR doll*" or PractiMan or Prestan or LifeSaveHer or Simulaids or "Life form" or "Nasco Healthcare" or Laerdal or WorldPoint or Simulaids or womanikin or "Resusci Anne") adj10 (diversity or diverse or variation* or represent* or sex or gender* or woman or women or female* or breast* or race or racial or ethnic* or minorit* or black* or asian* or hispanic* or "non white*" or "people of color" or "people of colour" or "person of color" or "person of colour" or POC or WOC or BIPOC or "skin color*" or "skin colour*" or age or infant* or newborn* or neonat* or baby or babies or child* or pediatr* or paediatr* or aged or elder* or geriatric* or "old person*" or "old people" or appearance* or "body shape*" or "body habit*" or "body build*" or "body type*" or somatotype* or obese or obesity or overweight* or bariatric* or "non lean" or pregnan* or socioeconomic* or sociodemographic* or demographic* or bias*)).ti,ab,hw,id. | 406 |
| 20 | 18 or 19 | 430 |
| 21 | 4 and 20 | **6** |

**Search History CINAHL conducted at 7th May 2024**

| **#** | **Search strategy** | **Results** |
| --- | --- | --- |
| S21 | S5 AND S20 | **268** |
| S20 | S18 OR S19 | 1,872 |
| S19 | TI ( ((manikin* or mannikin* or mannequin* or dummy or dummies or "simulated patient*" or "patient simulator*" or "CPR doll*" or PractiMan or Prestan or LifeSaveHer or Simulaids or "Life form" or "Nasco Healthcare" or Laerdal or WorldPoint or Simulaids or womanikin or "Resusci Anne") N10 (diversity or diverse or variation* or represent* or sex or gender* or woman or women or female* or breast* or race or racial or ethnic* or minorit* or black* or asian* or hispanic* or "non white*" or "people of color" or "people of colour" or "person of color" or "person of colour" or POC or WOC or BIPOC or "skin color*" or "skin colour*" or age or infant* or newborn* or neonat* or baby or babies or child* or pediatr* or paediatr* or aged or elder* or geriatric* or "old person*" or "old people" or appearance* or "body shape*" or "body habit*" or "body build*" or "body type*" or somatotype* or obese or obesity or overweight* or bariatric* or "non lean" or pregnan* or socioeconomic* or sociodemographic* or demographic* or bias*)) ) OR AB ( ((manikin* or mannikin* or mannequin* or dummy or dummies or "simulated patient*" or "patient simulator*" or "CPR doll*" or PractiMan or Prestan or LifeSaveHer or Simulaids or "Life form" or "Nasco Healthcare" or Laerdal or WorldPoint or Simulaids or womanikin or "Resusci Anne") N10 (diversity or diverse or variation* or represent* or sex or gender* or woman or women or female* or breast* or race or racial or ethnic* or minorit* or black* or asian* or hispanic* or "non white*" or "people of color" or "people of colour" or "person of color" or "person of colour" or POC or WOC or BIPOC or "skin color*" or "skin colour*" or age or infant* or newborn* or neonat* or baby or babies or child* or pediatr* or paediatr* or aged or elder* or geriatric* or "old person*" or "old people" or appearance* or "body shape*" or "body habit*" or "body build*" or "body type*" or somatotype* or obese or obesity or overweight* or bariatric* or "non lean" or pregnan* or socioeconomic* or sociodemographic* or demographic* or bias*)) ) | 879 |
| S18 | S16 AND S17 | 1,180 |
| S17 | (MH "Models, Anatomic") | 7,804 |
| S16 | S6 OR S7 OR S8 OR S9 OR S10 OR S11 OR S12 OR S13 OR S14 OR S15 | 1,877,259 |
| S15 | (MH "Sociodemographic Factors") | 11,380 |
| S14 | (MH "Somatotypes") | 329 |
| S13 | (MH "Aged+") | 955,781 |
| S12 | (MH "Child+") | 760,322 |
| S11 | (MH "Infant+") | 287,052 |
| S10 | (MH "Race Factors") | 32,544 |
| S9 | (MH "Ethnic Groups+") | 176,429 |
| S8 | (MH "Sex Factors") | 136,889 |
| S7 | (MH "Cultural Diversity+") | 18,377 |
| S6 | (MH "Health Inequities") | 2,291 |
| S5 | S1 OR S2 OR S3 OR S4 | 36,621 |
| S4 | TI ( ("Cardiopulmonary Resuscitation*" or "Cardio pulmonary Resuscitation*" or "cardiopulmonary reanimation*" or "cardio pulmonary reanimation*" or CPR or "basic life support*" or "cardiac life support*" or BLS or "chest compression*" or "cardiac massage*" or "heart massage*" or "mouth to mouth Resuscitation*" or defibrillator* or AED) ) OR AB ( ("Cardiopulmonary Resuscitation*" or "Cardio pulmonary Resuscitation*" or "cardiopulmonary reanimation*" or "cardio pulmonary reanimation*" or CPR or "basic life support*" or "cardiac life support*" or BLS or "chest compression*" or "cardiac massage*" or "heart massage*" or "mouth to mouth Resuscitation*" or defibrillator* or AED) ) | 27,339 |
| S3 | (MH "Defibrillators, Automated External") | 1,241 |
| S2 | (MH "Defibrillators") | 1,630 |
| S1 | (MH "Resuscitation, Cardiopulmonary+") | 17,312 |

**Search History Scopus conducted at 7th May 2024**

| **#** | **Query** | **Results** |
| --- | --- | --- |
| 1 | TITLE-ABS-KEY("Cardiopulmonary Resuscitation*" or "Cardio pulmonary Resuscitation*" or "cardiopulmonary reanimation*" or "cardio pulmonary reanimation*" or CPR or "basic life support*" or "cardiac life support*" or BLS or "chest compression*" or "cardiac massage*" or "heart massage*" or "mouth to mouth Resuscitation*" or defibrillator* or AED) | 109,333 |
| 2 | TITLE-ABS-KEY(((manikin* or mannikin* or mannequin* or dummy or dummies or "simulated patient*" or "patient simulator*" or "CPR doll*" or PractiMan or Prestan or LifeSaveHer or Simulaids or "Life form" or "Nasco Healthcare" or Laerdal or WorldPoint or Simulaids or womanikin or "Resusci Anne") W/10 (diversity or diverse or variation* or represent* or sex or gender* or woman or women or female* or breast* or race or racial or ethnic* or minorit* or black* or asian* or hispanic* or "non white*" or "people of color" or "people of colour" or "person of color" or "person of colour" or POC or WOC or BIPOC or "skin color*" or "skin colour*" or age or infant* or newborn* or neonat* or baby or babies or child* or pediatr* or paediatr* or aged or elder* or geriatric* or "old person*" or "old people" or appearance* or "body shape*" or "body habit*" or "body build*" or "body type*" or somatotype* or obese or obesity or overweight* or bariatric* or "non lean" or pregnan* or socioeconomic* or sociodemographic* or demographic* or bias*))) | 6,477 |
| 3 | 1 AND 2 | **351** |

**Search History Web of Science conducted at 7th May 2024**

| **#** | **Search Query** | **Results** |
| --- | --- | --- |
| 1 | TS=("Cardiopulmonary Resuscitation*" or "Cardio pulmonary Resuscitation*" or "cardiopulmonary reanimation*" or "cardio pulmonary reanimation*" or CPR or "basic life support*" or "cardiac life support*" or BLS or "chest compression*" or "cardiac massage*" or "heart massage*" or "mouth to mouth Resuscitation*" or defibrillator* or AED) | 85393 |
| 2 | TS=((manikin* or mannikin* or mannequin* or dummy or dummies or "simulated patient*" or "patient simulator*" or "CPR doll*" or PractiMan or Prestan or LifeSaveHer or Simulaids or "Life form" or "Nasco Healthcare" or Laerdal or WorldPoint or Simulaids or womanikin or "Resusci Anne") NEAR/10 (diversity or diverse or variation* or represent* or sex or gender* or woman or women or female* or breast* or race or racial or ethnic* or minorit* or black* or asian* or hispanic* or "non white*" or "people of color" or "people of colour" or "person of color" or "person of colour" or POC or WOC or BIPOC or "skin color*" or "skin colour*" or age or infant* or newborn* or neonat* or baby or babies or child* or pediatr* or paediatr* or aged or elder* or geriatric* or "old person*" or "old people" or appearance* or "body shape*" or "body habit*" or "body build*" or "body type*" or somatotype* or obese or obesity or overweight* or bariatric* or "non lean" or pregnan* or socioeconomic* or sociodemographic* or demographic* or bias*)) | 3758 |
| 3 | #2 AND #1 | **300** |

**Search History Informit conducted at 7th May 2024**

| **#** | **Query (Search in: All Fields)** | **Results** |
| --- | --- | --- |
| 1 | "Cardiopulmonary Resuscitation" OR "Cardio pulmonary Resuscitation" OR "cardiopulmonary reanimation" OR "cardio pulmonary reanimation" OR CPR OR "basic life support" OR "cardiac life support" OR BLS OR "chest compression" OR “chest compressions” OR "cardiac massage" OR "heart massage" OR "mouth to mouth Resuscitation" OR defibrillator* OR AED | 614 |
| 2 | manikin* OR mannikin* OR mannequin* OR dummy OR dummies OR “simulated patient” OR “patient simulator” OR “simulated patients” OR “patient simulators” OR “CPR doll” OR “CPR dolls” OR practiman OR prestan OR lifesaveher OR simulaids OR “life form” OR “nasco healthcare” OR laerdal OR worldpoint OR simulaids OR womanikin OR “resusci anne” | 326 |
| 3 | diversity OR diverse OR variation* OR represent* OR sex OR gender* OR woman OR women OR female* OR breast* OR race OR racial OR ethnic* OR minorit* OR black* OR asian* OR hispanic* OR “non white” OR “non whites” OR “people of color” OR “people of colour” OR “person of color” OR “person of colour” OR POC OR WOC OR BIPOC OR “skin color” OR “skin colour” OR “skin colors” OR “skin colours” OR age OR infant* OR newborn* OR neonat* OR baby OR babies OR child* OR pediatr* OR paediatr* OR aged OR elder* OR geriatric* OR “old person” OR “old persons” OR “old people” OR appearance* OR “body shape” OR “body shapes” OR “body habitus” OR “body habituses” OR “body habiti” OR “body build” OR “body builds” OR “body type” OR “body types” OR somatotype* OR obese OR obesity OR overweight* OR bariatric* OR “non lean” OR pregnan* OR socioeconomic* OR sociodemographic* OR demographic* OR bias* | 286771 |
| 4 | #1 AND #2 AND #3 | **8** |

**Search History ERIC conducted at 8th May 2024**

| **#** | **Query (Search in: All Fields)** | **Results** |
| --- | --- | --- |
| 1 | "Cardiopulmonary Resuscitation" OR "Cardio pulmonary Resuscitation" OR "cardiopulmonary reanimation" OR "cardio pulmonary reanimation" OR CPR OR "basic life support" OR "cardiac life support" OR BLS OR "chest compression" OR "chest compressions" OR "cardiac massage" OR "heart massage" OR "mouth to mouth Resuscitation" OR defibrillator OR defibrillators OR AED | 4,274 |
| 2 | manikin OR manikins OR mannikin OR mannikins OR mannequin OR mannequins OR dummy OR dummies OR "simulated patient" OR "patient simulator" OR "simulated patients" OR "patient simulators" OR "CPR doll" OR "CPR dolls" OR practiman OR prestan OR lifesaveher OR simulaids OR "life form" OR "nasco healthcare" OR laerdal OR worldpoint OR simulaids OR womanikin OR "resusci anne" | 739 |
| 3 | #1 AND #2 | **22** |
